# Automatically Identifying Twitter Users for Interventions to Support Dementia Family Caregivers: Annotated Data Set and Benchmark Classification Models

**DOI:** 10.1101/2022.05.18.22275268

**Authors:** Ari Z. Klein, Arjun Magge, Karen O’Connor, Graciela Gonzalez-Hernandez

## Abstract

**Background:** More than 6 million people in the United States have Alzheimer’s disease and related dementias, receiving help from more than 11 million family or other informal caregivers. A range of traditional interventions have been developed to support family caregivers; however, most of them have not been implemented in practice and remain largely inaccessible. While recent studies have shown that family caregivers of people with dementia use Twitter to discuss their experiences, methods have not been developed to enable the use of Twitter for interventions.

**Objective:** The objective of this study was to develop an annotated data set and benchmark classification models for automatically identifying a cohort of Twitter users who have a family member with dementia.

**Methods:** Between May 4, 2021 and May 20, 2021, we collected 10,733 tweets, posted by 8846 users, that mention a dementia-related keyword, a linguistic marker that potentially indicates a diagnosis, and a select familial relationship. Three annotators annotated one random tweet per user to distinguish those that indicate having a family member with dementia from those that do not. We used the annotated tweets to train and evaluate deep neural network classifiers based on pretrained transformer models. To assess the scalability of our approach, we, then, deployed automatic classification on tweets that were continuously collected between May 4, 2021 and March 9, 2022.

**Results:** Inter-annotator agreement was 0.82 (Fleiss’ kappa). A classifier based on a BERT model pretrained on tweets achieved the highest F_1_-score of 0.962 (precision = 0.946, recall = 0.979) for the class of tweets indicating that the user has a family member with dementia. The classifier detected 128,838 tweets that indicate having a family member with dementia, posted by 74,290 users between May 4, 2021 and March 9, 2022—that is, approximately 7500 users per month.

**Conclusions:** Our annotated data set can be used to automatically identify Twitter users who have a family member with dementia, enabling the use of Twitter on a large scale to not only explore family caregivers’ experiences, but also directly target interventions at these users.

## Introduction

More than 6 million people in the United States have Alzheimer’s disease and related dementias, and the burden is projected to double by 2060 [1]. Alzheimer’s disease is the sixth leading cause of death in the United States [2], and only 8% of people with dementia do not receive help from family members or other informal care providers [3], amounting to more than 11 million family or other unpaid caregivers in 2020 [4]. Caregivers of people with dementia are impacted physically, cognitively, socially, mentally, and financially. For instance, compared with non-caregivers, they are more vulnerable to disease due to chronic stress [5] and have lower durations and quality of sleep [6]. Compared with non-dementia caregivers, they are more likely to experience a decline in cognition [7] and social network size [8]. They are also more likely to experience depression than non-caregivers [9] and non-dementia caregivers [10], and depressive symptoms in dementia caregivers are associated with increased health care use and costs [11]. In addition to the increased costs of their personal health care, family caregivers of people with dementia pay for much of the recipient’s total care costs, with the costs being significantly higher for people with dementia than without dementia [12].

A range of traditional interventions have been developed to support family caregivers of people with dementia [13]; however, most of them have not been implemented in practice and remain largely inaccessible [14]. Recent systematic reviews have concluded that internet-based interventions are valued by family caregivers of people with dementia for their easy access [15] and can have beneficial effects on caregivers’ health [16]. While recent studies [17-23] have shown that family caregivers of people with dementia use Twitter to discuss their experiences, as far as we know, methods have not been developed to enable the use of Twitter as a platform for internet-based interventions. Given that nearly one of every four adults in the United States uses Twitter [24], Twitter may present a novel opportunity to reach family caregivers on a large scale, such as through user-targeted advertisements providing information about dementia, caregiving, resources, or services. The objective of this study was to develop an annotated data set and benchmark classification models for automatically identifying a cohort of Twitter users who have a family member with dementia.

## Methods

### Data Collection and Annotation

Between May 4, 2021 and May 20, 2021, we collected 67,060 publicly available tweets from the Twitter Streaming Application Programming Interface (API) that are in English, are not retweets, and include both a dementia-related keyword (e.g., *dementia, youngdementia, #yod, #ftd, alzheimer’s, alz, alzheimersdisease, mild cognitive impairment*) and a linguistic marker that potentially indicates a diagnosis (e.g., *diagnosed, diagnosis, has, got, developed, with, from*). We, then, searched these tweets for references to select familial relationships (e.g., *mom, dad, grandma, grandpa, wife, husband, sister, brother, daughter, son*), identifying 10,733 (16%) of the 67,060 tweets. We randomly sampled one tweet per user—8846 (82%) of the 10,733 tweets—and developed annotation guidelines to help three annotators distinguish tweets that indicate having a family member with dementia from those that do not.

### Automatic Classification

We performed benchmark supervised machine learning experiments to assess the utility of the annotated data set for automatically identifying Twitter users who have a family member with dementia. We used six deep neural network classifiers based on bidirectional encoder representations from transformers (BERT): the BERT-Base-Uncased [25], DistilBERT-Base-Uncased [26], RoBERTa-Large [27], BioBERT-Large-Cased [28], Bio+ClinicalBERT [29], and BERTweet-Large [30] pretrained models in the *Flair* Python library. We split the 8846 tweets into 80% (7077 tweets) and 20% (1769 tweets) random sets as training data and held-out test data, respectively, stratified based on the distribution of the binary annotated classes. Prior to automatic classification, we preprocessed the tweets by normalizing URLs and usernames, and lowercasing the text. For training, we used stochastic gradient descent (SGD) optimization, a batch size of 8, 15 epochs, and a learning rate of 0.001. During training, we fine-tuned all layers of the transformer model with our annotated tweets. To optimize performance, the model was evaluated after each epoch, on a 5% split of the training set. To assess the scalability of our approach, we, then, deployed automatic classification on 198,674 tweets, posted by 119,640 users, that were continuously collected from the Twitter Streaming API between May 4, 2021 and March 9, 2022 and mention a select familial relationship. To assess the potential for tailoring interventions, we used Carmen [31] to infer the geolocation of users that the classifier determined have a family member with dementia.

## Results

Among the 8846 annotated tweets, 8346 (94%) were dual annotated, and 500 (6%) were annotated by all three annotators. Inter-annotator agreement, based on the 500 tweets annotated by all three annotators, was 0.82 (Fleiss’ kappa). Upon resolving the disagreements, it was determined that 5946 (67%) of the tweets indicate that the user has a family member with dementia (“positive” class), and 2900 (33%) of the tweets do not (“negative” class). Table 1 presents the precision, recall, and F_1_-scores of six deep neural network classifiers for the “positive” class, evaluated on a held-out test set of 1769 (20%) of the 8846 annotated tweets. The classifier based on a model pretrained on tweets (BERTweet-Large) achieved the highest F_1_-score: 0.962 (precision = 0.946, recall = 0.979). The BERTweet classifier detected 128,838 tweets indicating that the user has a family member with dementia, posted by 74,290 users between May 4, 2021 and March 9, 2022—that is, approximately 7500 users per month. Carmen [31] inferred the geolocation of 31,653 (43%) of these 74,290 users.

**Table 1.** Precision, recall, and F_1_-scores of classifiers for detecting tweets indicating that the user has a family member with dementia.

| Classifier | Precision | Recall | $F_1$-score |
| --- | --- | --- | --- |
| BERT-Base-Uncased | 0.924 | 0.954 | 0.938 |
| DistilBERT-Base-Uncased | 0.930 | 0.942 | 0.936 |
| RoBERTa-Large | 0.918 | **0.982** | 0.949 |
| BioBERT-Large-Cased | 0.907 | 0.978 | 0.941 |
| Bio+ClinicalBERT | 0.903 | 0.958 | 0.930 |
| BERTweet-Large | **0.946** | 0.979 | **0.962** |

Table 2 presents examples of false positives and false negatives of the BERTweet classifier in the test set. Among the 68 false positives, 36 (47%) refer to people with dementia who are not or may not be select family members (Tweet1), 8 (12%) report that a family member has a condition other than dementia (Tweet 2), and 5 (7%) merely speculate that a family member has dementia (Tweet 3). Another 8 (12%) of the 68 false positives were a result of manual annotation errors. Among the 25 false negatives, 14 (56%) use deixis or anaphora, requiring additional context in the tweet to understand that a non-first-person determiner (e.g., *their* in Tweet 4) actually refers to the user, or that a personal pronoun (e.g., *she* in Tweet 5) refers to a select family member with dementia. Furthermore, 12 (86%) of these 14 tweets also include references to people who are not family members or do not have dementia. Another 4 (16%) of the 25 false negatives were a result of manual annotation errors.

**Table 2.** Sample false positives and false negatives of a BERTweet classifier for detecting tweets indicating that the user has a select family member with dementia.

|  | **Tweet** | **Actual** | **Predicted** |
| --- | --- | --- | --- |
| 1 | Evelyn has dementia, I know. But when she asked me today how my dad was doing... it still hurt. | - | + |
| 2 | We really don't have a clue about what causes Alzheimer's. We don't have a clue about Parkinson's, which is what got my dad, either. | - | + |
| 3 | I just listened to the Everywhere at The End of Time, by The Caretaker, and thought about my grandmother. The songs are about dementia, something my grandma wasn't clearly diagnosed with, but it hit hard. | - | + |
| 4 | If someone tells u their parent has Alzheimer's please don't say your grandparent or great aunt did too. I appreciate that u can relate to the experience but it is so different. Tell me a different time. | + | - |
| 5 | I have a family member who is vulnerable and two children in their late 20s. I didn't want to risk passing virus to her or from her to my family member. My sister made a bubble with her and her carers. She has dementia so she probably hasn't missed me! | + | - |

## Discussion

The benchmark performance of automatic classification demonstrates that our annotated data set has utility for accurately identifying Twitter users who have a family member with dementia, and deploying automatic classification on unlabeled tweets demonstrates that a large cohort of users can be identified. Although we assumed that having “close” relatives with dementia would more likely imply the users’ involvement in caregiving, the users identified in this study may not be caregivers or may have been caregivers but are no longer. Nonetheless, we believe that limiting our identification of caregivers to users who explicitly state that they are providing ongoing care would underutilize the potential of Twitter for scaling up accessible interventions. Furthermore, our approach to identifying users and our ability to identify the geolocation of nearly half of them would enable opportunities to tailor interventions based on the care recipient and community.

## Conclusions

This paper presented an annotated data set and benchmark classification models for automatically identifying Twitter users who have a family member with dementia, enabling the use of Twitter on a large scale to not only explore family caregivers’ experiences among their tweets, but also directly target interventions at these users.

## Data Availability

All data produced in the present study are available upon reasonable request to the authors.

## Acknowledgments

AZK designed the data collection, edited the annotation guidelines, conducted the error analysis, and wrote the manuscript. AM performed the machine learning experiments, deployed the BERTweet classifier, and edited the manuscript. KO developed the annotation guidelines, annotated the Twitter data, and edited the manuscript. GGH conceptualized the study and edited the manuscript. The authors thank Ivan Flores for contributing to software applications, and Alexis Upshur and Aiden McRobbie-Johnson for contributing to annotating the Twitter data. This work was supported by the National Library of Medicine (grant number R01LM011176).

